# Expected impact of MRI-targeted biopsy interreader variability among uropathologists on ProScreen prostate cancer screening trial: a pre-trial validation study

**DOI:** 10.1101/2023.08.08.23293780

**Authors:** Ronja Hietikko, Tuomas Mirtti, Tuomas P. Kilpeläinen, Teemu Tolonen, Anne Räisänen-Sokolowski, Stig Nordling, Jill Hannus, Marita Laurila, Kimmo Taari, Teuvo LJ Tammela, Reija Autio, Kari Natunen, Anssi Auvinen, Antti Rannikko

## Abstract

**Background:** Prostate cancer (PCa) histology, particularly the Gleason score, is an independent prognostic predictor in PCa. Little is known about the inter-reader variability in grading of targeted prostate biopsy based on magnetic resonance imaging (MRI).

**Objective:** To assess inter-reader variability in Gleason grading of MRI-targeted biopsy among uropathologists and its potential impact on a population-based randomized PCa screening trial (ProScreen).

**Design, setting, and participants:** From June 2014 to May 2018, 100 men with clinically suspected PCa were retrospectively selected. All men underwent prostate MRI and 86 underwent targeted prostate of the prostate.

**Intervention:** Six pathologists individually reviewed the pathology slides of the prostate biopsies.

**Outcome measurements and statistical analysis:** The five-tier ISUP (The International Society of Urological Pathology) grade grouping (GG) system was used. Fleiss’ weighted kappa (κ) and Model based kappa for associations were computed to estimate the combined agreement between individual pathologists.

**Results and limitations:** GG reporting of targeted prostate was highly consistent among the trial pathologists. Inter-reader agreement for cancer (GG 1-5) vs. benign was excellent (Model-based kappa 0.90, Fleiss’ kappa κ = 0.90) and for clinically significant prostate cancer (csPCa) (GG 2-5 vs. GG 0 vs GG1) it was good (Model-based kappa 0.70, Fleiss’ kappa κ 0.67).

**Conclusions:** Inter-reader agreement in grading of MRI-targeted biopsy was good to excellent, while it was fair to moderate for MRI in the same cohort, as previously shown. Importantly, there was wide consensus by pathologists in assigning the contemporary GG on MRI-targeted biopsy suggesting high reproducibility of pathology reporting in the ProScreen trial.

**Patient summary:** It is currently unknown to what extent pathologists differ in their evaluation of histopathology in MRI-targeted prostate biopsies. We show that the agreement is good to excellent. We expect individual pathologist to have a minimal impact on MRI-based prostate cancer screening including the ProScreen trial.

## 1. Introduction

Population-based prostate cancer (PCa) screening using prostate-specific antigen (PSA) and standard transrectal ultrasound-guided prostate biopsies in men with elevated PSA levels reduces cancer-specific mortality ^1^. However, such screening also results in substantial overdiagnosis and overtreatment of clinically insignificant prostate cancer (cisPCa) ^1 2^.

Multiparametric magnetic resonance imaging (mpMRI) of the prostate and subsequent targeted prostate biopsies of identified lesions with clinical suspicion of PCa (PI-RADS 3-5) are a promising diagnostic pathway ^3^. In studies involving men with a suspected PCa, mpMRI improves the detection of csPCa and decreases cisPCa diagnosis compared to systematic biopsies ^3^. A recent study by Eklund and colleagues showed that a pre-biopsy MRI only was not inferior to systematic biopsies for detecting csPCa (21% compared to 18%), while detection of cisPCa was reduced by two-thirds^4^.

We initiated a population-based, prospective randomized PCa screening trial (ProScreen) in 2018. Unlike the STHLM3MRI and the Göteborg-2 studies, ProScreen trial is powered to evaluate PCa mortality as the primary endpoint ^4 5^. In the ProScreen trial, screen-positive men are referred to mpMRI with targeted prostate of the MRI visible lesion(s) only ^6^. Thus, the emphasis is on minimizing overdiagnosis, while retaining the previously established PCa mortality reduction from screening. To this end, correct identification of csPCa by the pathologists is important for proper treatment selection. Further, to our best knowledge, no previous studies have been published on interobserver agreement of pathologists’ interpretation of MRI-targeted prostate biopsies. Importantly, the last ISUP consensus conference emphasized the differences between reporting of systematic biopsies and targeted prostate ^7^. Therefore, the aim of this study was to evaluate MRI-targeted biopsy related interreader variability and its expected impact on the ProScreen trial.

## 2. Patients and methods

In the ProScreen trial a total of 67,000 men aged 50-63 years were identified from the population registry and randomized to either a screening arm or a control arm in a 1:3 ratio ^6^. The men in the control arm were not contacted. The men in the screening arm were invited to a screening test. A PSA ≥ 3.0 ng/ml triggered the next stage of screening, i.e. the four kallikrein (4Kscore) test ^8^. Men with a 4Kscore ≥ 7.5% were referred for MRI. Men with PI-RADS 3-5 lesion in MRI are then invited for fusion biopsies of the target lesions only. Of the men with negative MRI, only those with PSA density ≥ 0.15 ng/ml/ml are invited for systematic 12-core biopsies.

Here, we chose a cohort of 100 men who had been referred to the Helsinki University Hospital (HUS) for suspected PCa before the ProScreen trial. Men had varying baseline risk for PCa. The aim was to evaluate interreader variability in MRI and MRI-targeted biopsy. All 100 men were included in the previously reported study of interreader variability in MRI, and the cohort selection and patient demographics have been reported earlier ^9^. For this study, 91 men had undergone MRI before diagnostic biopsies, whereas for nine men, the MRI was used post-biopsy in cancer staging before definitive treatment. The biopsies were taken between June 2014 and May 2018 using MRI-fusion technique (UroNav, Philips, The Netherlands) to perform transrectal sampling of 2 to 4 biopsy cores per suspicious region of interest (ROI). Six patients’ samples could not be processed and viewed with cloud viewer due technical issues and were excluded from the final analysis.

All hematoxylin and eosin-stained glass slides of the 85 biopsies were included, representing the full spectrum of Gleason scores and no preselection of any kind was made. Slides were pseudonymized and digitally scanned using Pannoramic Flash III slide scanner (3D Histec, Budapest, Hungary) with a pixel resolution of 0.26um/pixel and reviewed with Aiforia cloud viewer software (Aiforia Technologies, Helsinki, Finland).

Six urological pathologists reviewed the slides and filled out a structured pathological assessment query including the number of glasses and biopsies, the length of biopsies and carcinoma, percent of Gleason pattern 4 or 5 and total ISUP Grade Group. The pathology reports of the primary ROI (ROI1), determined by the radiologist’s selection on the most significant ROI, were further analyzed. Clinical experience of the pathologists varied from three to fifty years (median 12.5, IQR 5.2-35.0). The pathologists were unaware of the other data regarding the patients. The original diagnostic pathology report on biopsies of ROI1 was collected.

### 2.1 Statistical analysis

We analyzed the agreement between all pathologists using model-based kappa for association, which is the preferred method when there are more than two raters and the classifications are ordinal ^11^.

Model-based kappa for association consider not only the exact agreement, but also the ratings close to each other, and the kappa value is computed giving weights to the classifications. Higher weights are given for the categories that are close to each other ^10^.

Along Model-based kappa, we have also used Fleiss kappa for comparison. Fleiss kappa is well known and more commonly used, but can be only used for categorical, not ordinal data. Fleiss kappa values and the Model based kappa for associations were computed with R using *irr* and *modelkappa* package. The interobserver agreement for grade groups are illustrated using R package *superheat*. Further the biopsies are clustered with k-means clustering into three groups, while the observers are clustered with hierarchical clustering using Euclidean distance and complete linkage. This retrospective analysis was evaluated and approved by the HUS (HUS/333/2019).

## 3. Results

The median age of the study participants at biopsy was 68.8 years (interquartile range [IQR] 60.9-75.0) and the median PSA was 9.1 ng/ml (IQR 6.7-13.8). The median number of biopsies obtained from the index lesion (ROI1) was 2.5 (IQR 2.0-3.0). The reported median length of an individual biopsy was 11.9 mm (IQR 10.4-13.5) and the median length of the cancer in a particular biopsy was 4.9 mm (IQR 3.0-7.8) (Table 1).

**Table 1.**
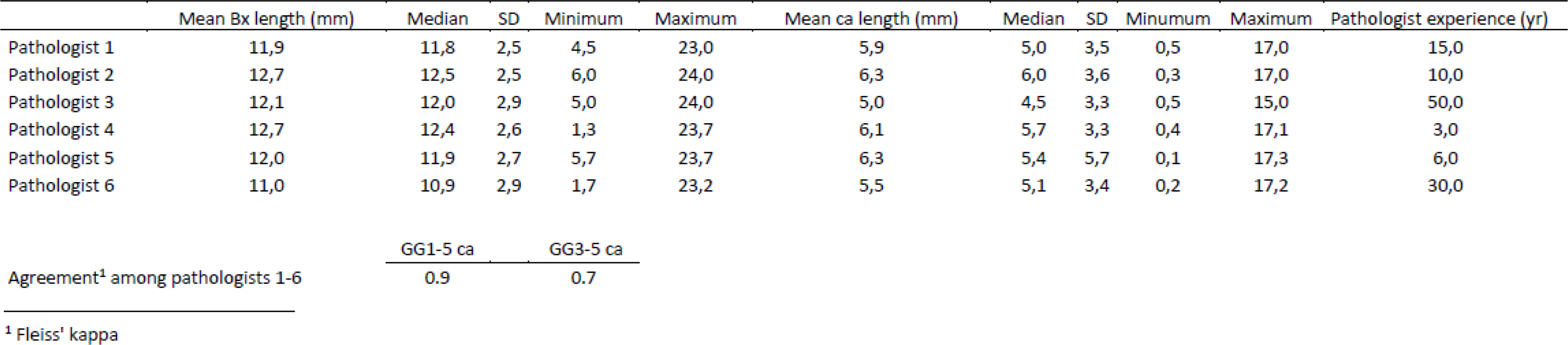
Index lesion-wise comparison of pathological characteristics and agreement on aggregated GG between study pathologists.

|  | Mean Bx length (mm) | Median | SD | Minimum | Maximum | Mean ca length (mm) | Median | SD | Minimum | Maximum | Pathologist experience (yr) |
| --- | --- | --- | --- | --- | --- | --- | --- | --- | --- | --- | --- |
| Pathologist 1 | 11,9 | 11,8 | 2,5 | 4,5 | 23,0 | 5,9 | 5,0 | 3,5 | 0,5 | 17,0 | 15,0 |
| Pathologist 2 | 12,7 | 12,5 | 2,5 | 6,0 | 24,0 | 6,3 | 6,0 | 3,6 | 0,3 | 17,0 | 10,0 |
| Pathologist 3 | 12,1 | 12,0 | 2,9 | 5,0 | 24,0 | 5,0 | 4,5 | 3,3 | 0,5 | 15,0 | 50,0 |
| Pathologist 4 | 12,7 | 12,4 | 2,6 | 1,3 | 23,7 | 6,1 | 5,7 | 3,3 | 0,4 | 17,1 | 3,0 |
| Pathologist 5 | 12,0 | 11,9 | 2,7 | 5,7 | 23,7 | 6,3 | 5,4 | 5,7 | 0,1 | 17,3 | 6,0 |
| Pathologist 6 | 11,0 | 10,9 | 2,9 | 1,7 | 23,2 | 5,5 | 5,1 | 3,4 | 0,2 | 17,2 | 30,0 |

Agreement<sup>1</sup> among pathologists 1-6
|  |  |
| --- | --- |
| GG1-5 ca | GG3-5 ca |
| 0.9 | 0.7 |

<sup>1</sup> Fleiss' kappa

The distributions of the assigned grades for each case by the study pathologists are shown in Table 2. In the original diagnostic pathology reports, 69 men were diagnosed with PCa. Of them, 10 (11.8%) had GG 1 cancer, 26 (29.4%) GG 2 cancer, 18 (20.0%) GG 3, 8 (11.8%) GG 4 and 7 (8.2%) GG 5 cancer in the biopsies. The proportion of biopsies reported as benign by the pathologists ranged from 18.6% to 23.3%. The detection of csPCa ranged from 44.7% to 77.6%. The patient-level GG assessments of all the observers grouped by the original clinical pathology report are illustrated in Figure 1.

**Table 2.** The distribution of the highest ISUPgrade 1-5 defined for all patientsfrom index lesion.

| Proposed grade | Clinical | Pathologist 1 | Pathologist 2 | Pathologist 3 | Pathologist 4 | Pathologist 5 | Pathologist 6 | All pathologists |
| --- | --- | --- | --- | --- | --- | --- | --- | --- |
| GG0 | 20 % | 22 % | 20 % | 20 % | 19 % | 20 % | 18 % | 20 % |
| GG1 | 11 % | 16 % | 22 % | 35 % | 4 % | 31 % | 21 % | 22 % |
| GG2 | 29 % | 15 % | 17 % | 6 % | 20 % | 16 % | 20 % | 16 % |
| GG3 | 21 % | 19 % | 12 % | 8 % | 18 % | 14 % | 11 % | 14 % |
| GG4 | 11 % | 24 % | 15 % | 26 % | 22 % | 11 % | 11 % | 18 % |
| GG5 | 8 % | 4 % | 14 % | 5 % | 18 % | 8 % | 20 % | 11 % |
| GG1-5 | 80 % | 78 % | 80 % | 80 % | 81 % | 80 % | 82 % | 80 % |
| GG2-5 | 69 % | 61 % | 58 % | 45 % | 78 % | 49 % | 61 % | 59 % |
| GG3-5 | 40 % | 46 % | 41 % | 39 % | 58 % | 33 % | 41 % | 43 % |

**Fig. 1.**
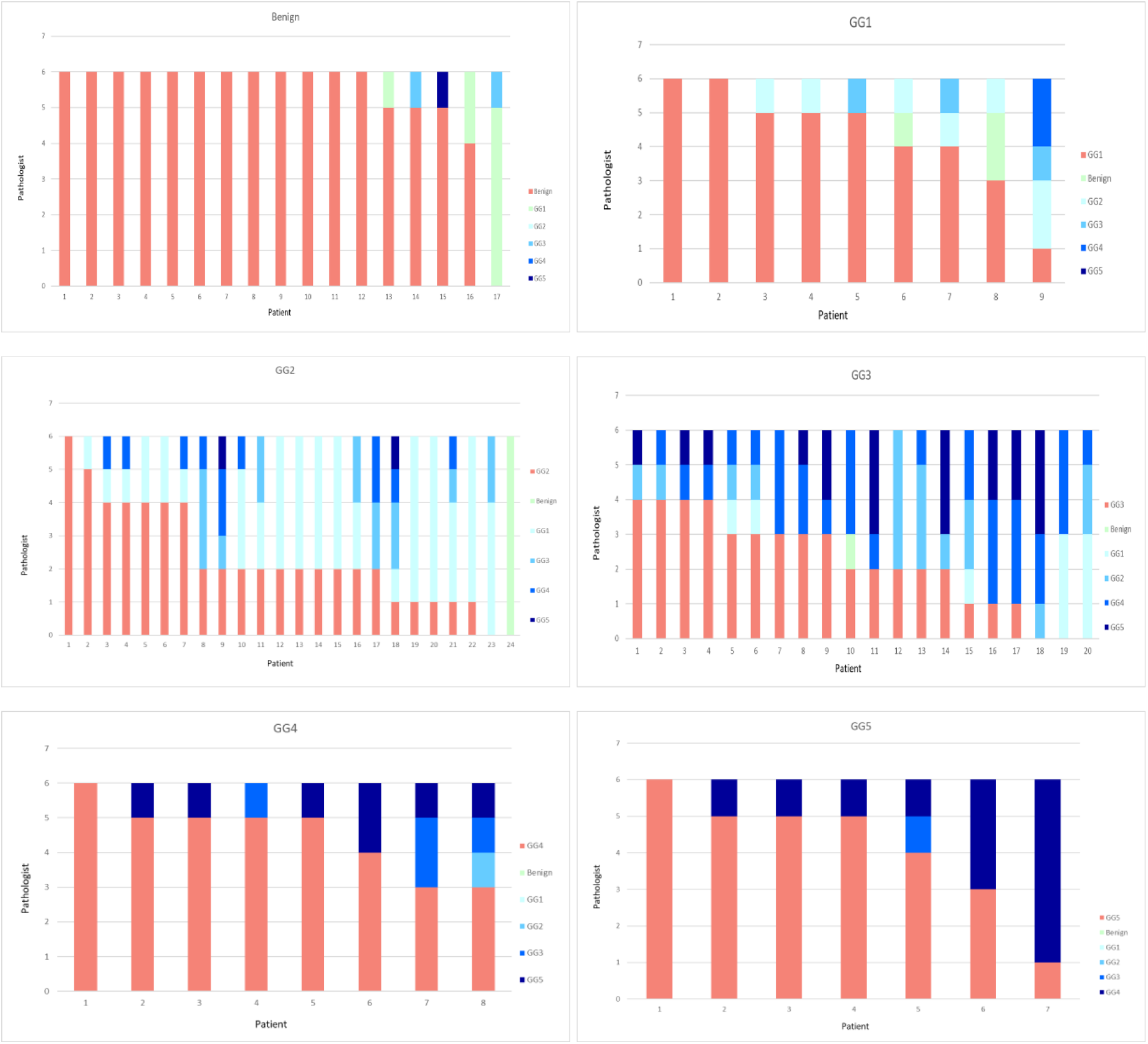
The number of pathologists identifying Gleason grade in prostate biopsies grouped by original pathological result.

We found complete agreement on the GG among all (6 /6) pathologists in 18 of 85 (21.2%) cases. Of the 18 cases with complete agreement, 72.2% (13 /18) were benign. We defined the consensus level as at least 2/3 agreement among the pathologists for a case according to the practice in all consensus meetings organized under the auspices of ISUP during the past decade ^11^. With this criterion, consensus was reached for 65.9% (56 /85) of the cases. The distribution of 2/3 grading consensus for ISUP GGs is shown in Table 3. The consensus grade differed from the initial grading in 13 cases. Almost all (92.3%) of these cases were in agreement within ±1 of the consensus GG.

**Table 3.**
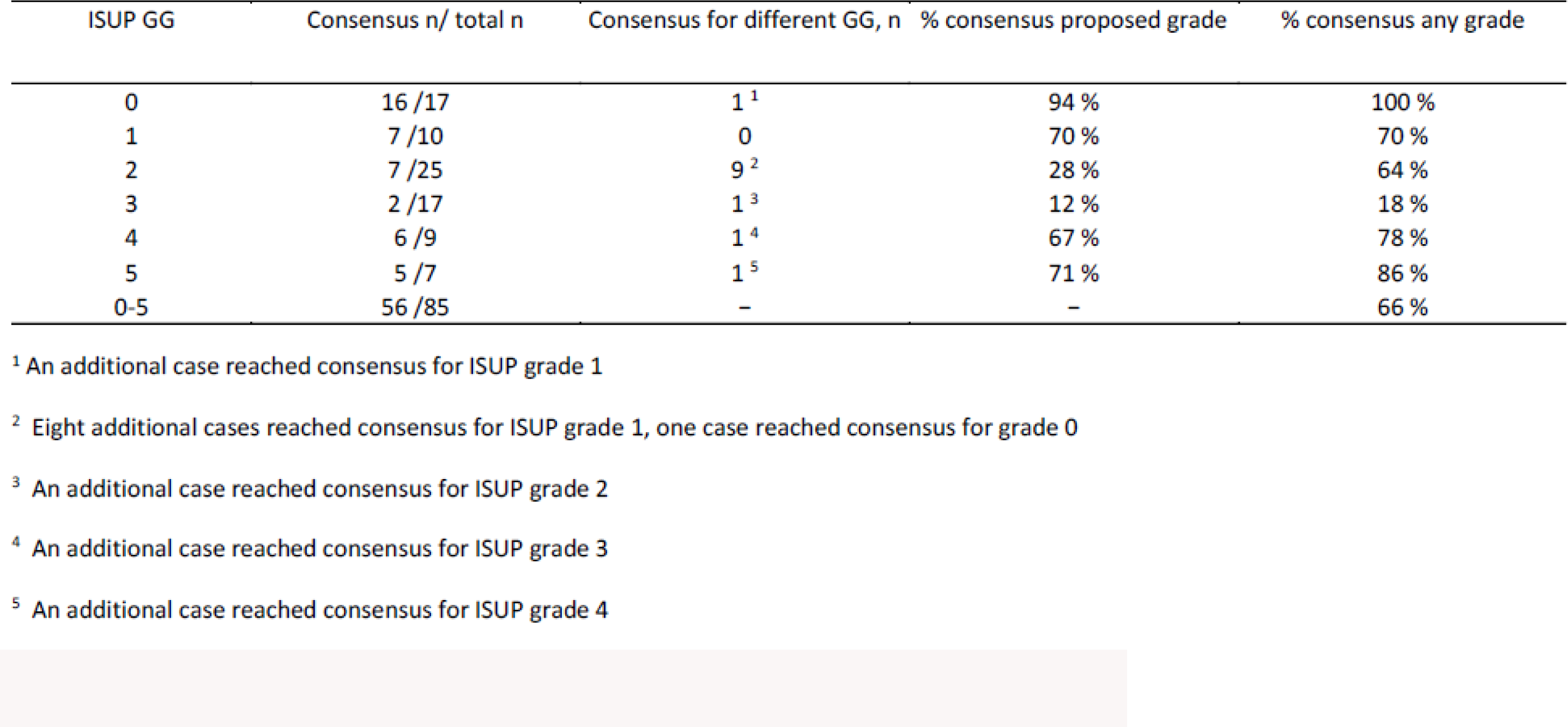
Reproducibility by proposed ISUPgrade among all cases with the consensus level defined as at least 2/3 of all pathologist.

| ISUP GG | Consensus n/ total n | Consensus for different GG, n | % consensus proposed grade | % consensus any grade |
| --- | --- | --- | --- | --- |
| 0 | 16/17 | 1<sup>1</sup> | 94 % | 100 % |
| 1 | 7/10 | 0 | 70 % | 70 % |
| 2 | 7/25 | 9<sup>2</sup> | 28 % | 64 % |
| 3 | 2/17 | 1<sup>3</sup> | 12 % | 18 % |
| 4 | 6/9 | 1<sup>4</sup> | 67 % | 78 % |
| 5 | 5/7 | 1<sup>5</sup> | 71 % | 86 % |
| 0-5 | 56/85 | - | - | 66 % |

<sup>1</sup> An additional case reached consensus for ISUP grade 1
<sup>2</sup> Eight additional cases reached consensus for ISUP grade 1, one case reached consensus for grade 0
<sup>3</sup> An additional case reached consensus for ISUP grade 2
<sup>4</sup> An additional case reached consensus for ISUP grade 3
<sup>5</sup> An additional case reached consensus for ISUP grade 4

No consensus was reached (agreement among pathologists below 2/3) for 34.1% (29 /85) of the cases. The most common source of disagreement was the estimated proportion of Gleason patterns 3 and 4. This reflects the challenges in distinguishing GG 2 from GG 3, as seen in six cases (21.4%). The agreement among the observers in a comparison including all six categories of cancer and benign was good (Model-based kappa 0.65, 95% CI 0.59 - 0.70). Agreement among pathologists for cancer (GG 1-5) vs. benign was excellent (Model-based kappa 0.90, Fleiss’ kappa κ = 0.90). For three-category comparison between csPCa (GG 2-5) vs. cisPCa (GG 1) vs. benign (GG 0) the inter-observer agreement was good (Model-based kappa 0.70, Fleiss’ kappa 0.67). The heatmap visualization shows the interobserver agreement for grade groups (Figure 2).

**Fig. 2.**
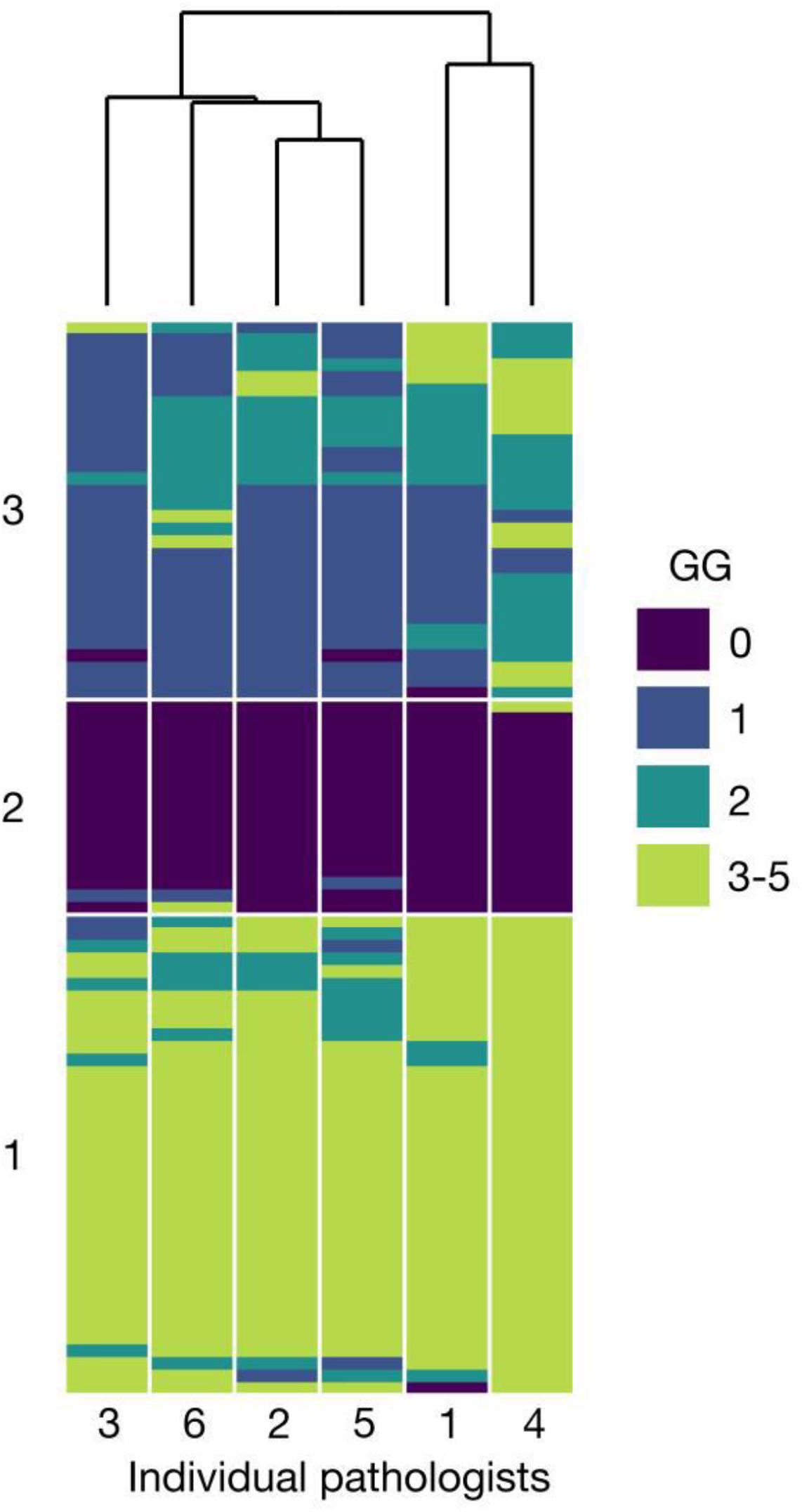
Heatmap visualization of the interobserver agreement for grade group. Individual pathologist are on the x-axis, colors represent grade groups for each ROI1 biopsy (GG 0 =benign). Pathologists and biopsies are ordered based on their similarity resulting from cluster analysis.

## 4. Discussion

Most guidelines for PCa screening in Europe and North America do not recommend organized population-wide screening ^12 13^. Despite a recent recommendation by the European Commission to implement PCa in national screening programs, there is no high-quality scientific evidence from randomized screening trials confirming that MRI-based, or a novel biomarker-based, PCa screening would decrease mortality from prostate cancer.

Histological grading is one of the most important prognostic factors of PCa because of its validated prediction of the clinical behavior of cancer ^14 15^. Interestingly, previous screening trials have not assessed the diagnostic agreement of the pathological reporting prior to the study initiation ^1 16^. Nor have multi-center diagnostic trials comparing MRI-targeted biopsies to systematic biopsies ^17 18^. Here, we show that the interreader agreement among pathologists was good to excellent in grading of MRI-targeted biopsies. Therefore, the expected impact of variability on the MRI-based ProScreen screening trial is minimal. Sufficient agreement between pathologists is crucial for maintaining the value of the Gleason grading system as a diagnostic and prognostic tool and in determining the appropriate treatment for a patient ^8^. According to some studies, 10-13% of PCa patients would receive different treatment recommendation after re-evaluation of biopsy specimen ^19 20 17^. Through correct assessment of GG, men can be properly assigned to either receive radical or more conservative treatment, to maximize the balance between the benefit and the harm.

In the benign vs. cancer comparison, the more commonly used Fleiss’ kappa, which better fits the two-category comparison, was similar to the model-based kappa (Fleiss’ kappa 0.90 vs. Model-based kappa 0.90). Overall, the model-based kappa is better suited for multi-categorical association analysis between several observers. Thus, it is not possible to directly compare our results with most of the previously published, systematic biopsy-based studies using Cohen’s or Fleiss’ kappa methodology. In addition, comparison across studies is challenging due to variation in definition of agreement, the type of investigated tissue (e.g., biopsies, radical prostatectomy specimens, transurethral resection specimens, a mixture of these, tissue microarray spots), different grouping of Gleason scores, the number of pathologists involved, and the number of specimens investigated. However, the agreement was better than in most other studies, in which kappa value has been calculated. The reported interobserver agreement among general pathologists for different comparisons has ranged from fair to moderate ^21 22 23^ although better results have also been reported ^24^. The reproducibility among uropathologists tends to be better than among the general pathologists, usually ranging with between kappa values 0.56 – 0.67 ^21 24 25 26^. We also noticed that the experience of the pathologist influences the results. Observer number four was an outlier in terms of years of experience. When we excluded this pathologist from the analysis, the agreement was higher among the more experienced pathologists (data not shown). This supports our practice in the ProScreen trial of uropathologists signing out the biopsies among the routine practice.

We observed the highest level of consensus in identifying cancers of any grade (GG 1-5). With more aggressive cancer (GG 4-5), the kappa value was lower than for lower grade tumors (GG 0-3), although still good. This probably reflects the challenge and variation in estimating the proportions between growth patterns, and possibly difficulties in detecting small foci of growth pattern 5.

Similar to our findings on MRI-related inter-reader variation ^9^, the extremes of the scale seem to be consistently reported, while the intermediate zone with borderline cases is challenging. We found the highest consensus with GG 0 (100.0%) and GG 5 (85.7%) and lower consensus within GG 3 (25.0%) and GG 2 (68.0%) cancer. In a PSA-based screening study, as many as 37.5% of biopsies were benign and only 8.0% of diagnosed cancers were GG 3, suggesting that the overall reproducibility in a screening may be even better though MRI targeting likely has an impact on this ^27^. The most common source of disagreement was separation of Gleason grade pattern 3 from pattern 4. The distinction between these two patterns was also recognized as a challenge in previous studies ^24 26^. Egevad et al. found a specific challenge in differentiating tangentially cut GG 1 from GG 2 for cases with poorly formed or fused glands ^28^. Further, fused glands or small glands without lumina may be interpreted as tangentially sectioned Gleason grade pattern 3 or as a focal Gleason grade pattern 4 ^29^. Zhou et al. reported that any case with ≤ 5 poorly formed glands should not be graded as Gleason pattern 4 ^30^. The ISUP 2014 revision of the Gleason grading system suggested that there should be more than occasional structures of this type for a tumor to qualify as Gleason pattern 4, otherwise they may represent tangential cuts. Previous studies have indicated that the reproducibility of Gleason pattern 4 with cribriform pattern is higher than Gleason pattern 4 with poorly formed or fused glands ^11 29^. All the above emphasize the importance of regular training, knowledge exchange between pathologists and intra-institutional peer evaluation, as making the decision on final GG is often subjective, especially in cases composed of Gleason grade patterns 3 and 4.

As the current clinical practice in diagnosing PCa relies heavily on targeted prostate alone or in conjunction with systematic biopsies, it is important that pathologists follow the guidelines for reporting ^7^. Our study is the first to evaluate the interobserver agreement of multiple pathologists on MRI-targeted diagnostic prostate biopsies. Assessing a lesion-wise aggregate GG has been shown to correlate better with RP GG than core-wise highest GG ^31^, which again emphasizes the need to adhere to reporting guidelines.

The present study has some inherent limitations. MRI-targeted biopsies were obtained from men with clinical suspicion for PCa, not from a screening cohort. This may influence the generalizability of the study results to a screening study with lower underlying PCa risk. However, our study was not designed to assess the diagnostic performance, thus the related limitations such as high prevalence of the disease and verification bias are not essential. Further, the aim was to investigate the interreader agreement among the pathologist for targeted prostate specifically. Therefore, we chose a study cohort with relatively even distribution of different histopathologies. Moreover, contrary to clinical routine, pathologists were not allowed to consult a colleague when faced with challenging cases. This, however, likely underestimates the interreader agreement observed, especially in more aggressive cancers. When extrapolating the study results on the ProScreen trial, these limitations should not have a major effect as agreement between benign and cisPCa versus csPCa was good, thus supporting clinical decision making on cancer treatments. Given that the same teams of pathologists will evaluate PCa cases in both the screening and control arms, among screening participants and non-participants, any variability in grading is likely to results in nondifferential misclassification, and hence it is expected to slightly decrease the differences between the compared groups.

## 5. Conclusion

The inter-reader agreement of MRI-targeted biopsy was good to excellent and better than the previously published inter-reader agreement for MRI from the same cohort. Therefore, it is plausible to assume that routine clinical histopathological evaluation is not likely to materially impact ProScreen trial results.

## Data Availability

All data produced in the present study are available upon reasonable request to the authors

## References

1. Hugosson J, Roobol MJ, Månsson M, et al. A 16-yr Follow-up of the European Randomized study of Screening for Prostate Cancer. Eur Urol. 2019;76(1):43–51. doi:10.1016/j.eururo.2019.02.009

2. Welch HG, Albertsen PC. Reconsidering Prostate Cancer Mortality - The Future of PSA Screening. N Engl J Med. 2020;382(16):1557–1563. doi:10.1056/NEJMms1914228

3. Drost FJH, Osses DF, Nieboer D, et al. Prostate MRI, with or without MRI-targeted biopsy, and systematic biopsy for detecting prostate cancer. Cochrane Database of Systematic Reviews. 2019;2019(4). doi:10.1002/14651858.CD012663.pub2

4. Nordström T, Discacciati A, Bergman M, et al. Prostate cancer screening using a combination of risk-prediction, MRI, and targeted prostate biopsies (STHLM3-MRI): a prospective, population-based, randomised, open-label, non-inferiority trial. Lancet Oncol. 2021;22(9):1240–1249. doi:10.1016/S1470-2045(21)00348-X

5. Kohestani K, Månsson M, Arnsrud Godtman R, et al. The GÖTEBORG prostate cancer screening 2 trial: a prospective, randomised, population-based prostate cancer screening trial with prostate-specific antigen testing followed by magnetic resonance imaging of the prostate. Scand J Urol. 2021;55(2):116–124. doi:10.1080/21681805.2021.1881612

6. Auvinen A, Rannikko A, Taari K, et al. A randomized trial of early detection of clinically significant prostate cancer (ProScreen): study design and rationale. Eur J Epidemiol. 2017;32(6):521–527. doi:10.1007/s10654-017-0292-5

7. van Leenders GJLH, van der Kwast TH, Grignon DJ, et al. The 2019 International Society of Urological Pathology (ISUP) Consensus Conference on Grading of Prostatic Carcinoma. American Journal of Surgical Pathology. 2020;44(8):e87–e99. doi:10.1097/PAS.0000000000001497

8. Bryant RJ, Sjoberg DD, Vickers AJ, et al. Predicting High-Grade Cancer at Ten-Core Prostate Biopsy Using Four Kallikrein Markers Measured in Blood in the ProtecT Study. JNCI: Journal of the National Cancer Institute. 2015;107(7). doi:10.1093/jnci/djv095

9. Hietikko R, Kilpeläinen TP, Kenttämies A, et al. Expected impact of MRI-related interreader variability on ProScreen prostate cancer screening trial: a pre-trial validation study. Cancer Imaging. 2020;20(1):72. doi:10.1186/s40644-020-00351-w

10. Nelson KP, Edwards D. Measures of agreement between many raters for ordinal classifications. Stat Med. 2015;34(23):3116–3132. doi:10.1002/sim.6546

11. Egevad L, Delahunt B, Berney DM, et al. Utility of Pathology Imagebase for standardisation of prostate cancer grading. Histopathology. 2018;73(1):8–18. doi:10.1111/his.13471

12. Parker C, Castro E, Fizazi K, et al. Prostate cancer: ESMO Clinical Practice Guidelines for diagnosis, treatment and follow-up. Annals of Oncology. 2020;31(9):1119–1134. doi:10.1016/j.annonc.2020.06.011

13. Mottet N, van den Bergh RCN, Briers E, et al. EAU-EANM-ESTRO-ESUR-SIOG Guidelines on Prostate Cancer—2020 Update. Part 1: Screening, Diagnosis, and Local Treatment with Curative Intent. Eur Urol. 2021;79(2):243–262. doi:10.1016/j.eururo.2020.09.042

14. Gleason DF. Classification of prostatic carcinomas. Cancer Chemother Rep. 1966;50(3):125–128.

15. Epstein JI, Egevad L, Amin MB, Delahunt B, Srigley JR, Humphrey PA. The 2014 International Society of Urological Pathology (ISUP) Consensus Conference on Gleason Grading of Prostatic Carcinoma. American Journal of Surgical Pathology. 2016;40(2):244–252. doi:10.1097/PAS.0000000000000530

16. Pinsky PF, Miller E, Prorok P, Grubb R, Crawford ED, Andriole G. Extended follow-up for prostate cancer incidence and mortality among participants in the Prostate, Lung, Colorectal and Ovarian randomized cancer screening trial. BJU Int. 2019;123(5):854–860. doi:10.1111/bju.14580

17. Kasivisvanathan V, Rannikko AS, Borghi M, et al. MRI-Targeted or Standard Biopsy for Prostate-Cancer Diagnosis. New England Journal of Medicine. 2018;378(19):1767–1777. doi:10.1056/NEJMoa1801993

18. Klotz L, Chin J, Black PC, et al. Comparison of Multiparametric Magnetic Resonance Imaging– Targeted Biopsy With Systematic Transrectal Ultrasonography Biopsy for Biopsy-Naive Men at Risk for Prostate Cancer. JAMA Oncol. 2021;7(4):534. doi:10.1001/jamaoncol.2020.7589

19. Nam RK, Oliver TK, Vickers AJ, et al. Prostate-Specific Antigen Test for Prostate Cancer Screening: American Society of Clinical Oncology Provisional Clinical Opinion. J Oncol Pract. 2012;8(5):315–317. doi:10.1200/JOP.2012.000715

20. Bibbins-Domingo K, Grossman DC, Curry SJ. The US Preventive Services Task Force 2017 Draft Recommendation Statement on Screening for Prostate Cancer. JAMA. 2017;317(19):1949. doi:10.1001/jama.2017.4413

21. Engers R. Reproducibility and reliability of tumor grading in urological neoplasms. World J Urol. 2007;25(6):595–605. doi:10.1007/s00345-007-0209-0

22. Ozkan TA, Eruyar AT, Cebeci OO, Memik O, Ozcan L, Kuskonmaz I. Interobserver variability in Gleason histological grading of prostate cancer. Scand J Urol. 2016;50(6):420–424. doi:10.1080/21681805.2016.1206619

23. Griffiths DFR, Melia J, McWilliam LJ, et al. A study of Gleason score interpretation in different groups of UK pathologists; techniques for improving reproducibility. Histopathology. 2006;48(6):655–662. doi:10.1111/j.1365-2559.2006.02394.x

24. Al Nemer AM, Elsharkawy T, Elshawarby M, Al-Tamimi D, Kussaibi H, Ahmed A. The updated grading system of prostate carcinoma: an inter-observer agreement study among general pathologists in an academic practice. APMIS. 2017;125(11):957–961. doi:10.1111/apm.12741

25. Thomsen FB, Marcussen N, Berg KD, et al. Repeated biopsies in patients with prostate cancer on active surveillance: clinical implications of interobserver variation in histopathological assessment. BJU Int. 2015;115(4):599–605. doi:10.1111/bju.12820

26. Allsbrook WC, Mangold KA, Johnson MH, et al. Interobserver reproducibility of Gleason grading of prostatic carcinoma: Urologic pathologists. Hum Pathol. 2001;32(1):74–80. doi:10.1053/hupa.2001.21134

27. Hugosson J, Månsson M, Wallström J, et al. Prostate Cancer Screening with PSA and MRI Followed by Targeted Biopsy Only. New England Journal of Medicine. 2022;387(23):2126–2137. doi:10.1056/NEJMoa2209454

28. Egevad L, Swanberg D, Delahunt B, et al. Identification of areas of grading difficulties in prostate cancer and comparison with artificial intelligence assisted grading. Virchows Archiv. 2020;477(6):777–786. doi:10.1007/s00428-020-02858-w

29. Kweldam CF, Nieboer D, Algaba F, et al. Gleason grade 4 prostate adenocarcinoma patterns: an interobserver agreement study among genitourinary pathologists. Histopathology. 2016;69(3):441–449. doi:10.1111/his.12976

30. Zhou M, Li J, Cheng L, et al. Diagnosis of “Poorly Formed Glands” Gleason Pattern 4 Prostatic Adenocarcinoma on Needle Biopsy. American Journal of Surgical Pathology. 2015;39(10):1331–1339. doi:10.1097/PAS.0000000000000457

31. Ren J, Melamed J, Taneja SS, et al. Prostate magnetic resonance imaging-targeted biopsy global grade correlates better than highest grade with prostatectomy grade. Prostate. 2023;83(4):323–330. doi:10.1002/pros.24464

